# The mobilome associated with Gram-negative bloodstream infections: A large-scale observational hybrid sequencing based study

**DOI:** 10.1101/2022.04.03.22273290

**Authors:** Samuel Lipworth, Willam Matlock, Liam Shaw, Karina-Doris Vihta, Gillian Rodger, Kevin Chau, Leanne Barker, Sophie George, James Kavanagh, Timothy Davies, Alison Vaughan, Monique Andersson, Katie Jeffery, Sarah Oakley, Marcus Morgan, Susan Hopkins, Timothy Peto, Derrick Crook, A. Sarah Walker, Nicole Stoesser

## Abstract

Plasmids carry genes conferring antimicrobial resistance (AMR), and other clinically important traits; their ability to move within and between species may provide the machinery for rapid dissemination of such genes. Existing studies using complete plasmid assemblies, which are essential for reliable inference, have been small and/or limited to those carrying particularly antimicrobial resistance genes (ARGs). In this study, we sequenced 1,880 complete plasmids from 738 isolates from bloodstream infections (BSI) in 2009 (194 isolates) and 2018 (368 isolates) in Oxfordshire, UK, plus a stratified selection from intervening years (176 isolates). We demonstrate that plasmids are largely, but not entirely, constrained to host species, although there is substantial overlap between species of plasmid gene-repertoire. Most ARGs are carried by a relatively small number of plasmid groups with biological features that are predictable. Plasmids carrying ARGs (including those encoding carbapenemases) share a putative ‘backbone’ of core genes with those carrying no such genes. These findings suggest that future surveillance should, in addition to tracking plasmids currently associated with clinically important genes, focus on identifying and monitoring the dissemination of high-risk plasmid groups with the potential to rapidly acquire and disseminate these genes.

## Introduction

Gram-negative bloodstream infections (BSI) are associated with substantial morbidity and mortality; their incidence continues to increase both in the UK and globally(1, 2). Multidrug-resistant and hypervirulent phenotypes are a particular concern, especially since genes conferring these characteristics (and others which may have either positive or negative fitness effects) are carried on plasmids, frequently in association with other smaller mobile genetic elements(3, 4). Plasmids are thought to facilitate the rapid dissemination of these genes within and between species. A detailed understanding of their biology and epidemiology is therefore likely to be crucial in tackling the global threat of antimicrobial resistance (AMR).

Complete and accurate genome assemblies, such as those produced by “hybrid” assemblies of short and long-read sequencing data, are crucial for the study of plasmid epidemiology. Until recently however, these have been prohibitively expensive for large-scale application and so whilst this approach has recently been used at scale to evaluate the plasmidome of environmental/agricultural isolates(5), to our knowledge its application to human-associated isolates has been mostly restricted to relatively small numbers of isolates selected based on AMR phenotype(3, 6). This phenotype-driven selection strategy has identified several plasmid types associated with the dissemination of key ARGs but it is currently not known whether similar plasmids are also found in susceptible populations. Recently, two studies have demonstrated the utility of network-based approaches to classify plasmid assemblies from public databases, offering insights into the host range of these plasmids, though such analyses suffer from sampling bias as well as a lack of clinical context and accurate metadata(7, 8). Therefore, the plasmidome associated with Gram-negative isolates causing both antimicrobial susceptible and sensitive clinical infections remains largely uncharacterised.

In this study, we generated a large collection of complete *E. coli*/*Klebsiella* spp. genomes from all BSIs collected in 2009 and 2018 in Oxfordshire, UK, as well as a representative sample from intervening years. Using this unprecedented dataset, we first sought to investigate the extent to which plasmids are shared and contribute to overlaps in the pangenome within and between species. We then sought to compare plasmids associated with ARG carriage to those that are not. Subsequently we investigated the dissemination dynamics of the most prevelant ESBL gene in the population, *bla*_*CTX-M-15*_, highlighting complex nested mobilisation that can only be unravelled using hybrid assembly. Finally we contextualised our findings by comparing our plasmid dataset to a large global collection and investigated whether features of “successful” plasmids and those with the potential for ARG carriage are predictable.

## Results

We successfully sequenced and assembled n=738 isolates of which 75% (553/738) were *E. coli* (n=153, 297, 103 in 2009, 2018, intervening years, respectively), 22% (161/738) *Klebsiella* spp. (n=39, 58, 64 in 2009, 2018, intervening years, respectively) and 3% (24/168) other Enterobacterales spp (details in Figure S1). In total, these isolates carried 1,880 plasmids with a median of 2 plasmids per isolate (interquartile range (IQR) 1-3). 10% (77/738) isolates carried none, 29% (211/738) carried one and 61% (450/738) more than one (Figure 1A). Of the n=661/738 isolates with at least one plasmid, 77% (508/661) carried at least one large plasmid (i.e., sequence length >100,000bp), and 94% 621/661) at least one large or medium plasmid (i.e., sequence length >10,000bp); of these 53% (329/621) also carried at least one small plasmid (i.e. sequence length <10,000bp). Carriage of one or more small plasmids in the absence of any medium or large plasmid was relatively rare at 6% (40/661). Rarefaction analysis suggested that a substantial number of plasmid groups remain unsampled and that there is a significantly greater diversity amongst groups containing smaller (<100,000bp) vs larger (≥ 100,000bp) plasmids (Figure 1B). There was some evidence that *Klebsiella* spp. isolates tended to carry slightly more plasmids than *E. coli*: median 2 (IQR 1-5) vs (median 2 (1-3) plasmids respectively (Kruskal-Wallis, p-value=0.03; Figure 1A), as did multi-drug resistant (MDR i.e., carriage of 3 ≥ ARG classes) vs. non-MDR isolates: (n=317/738 vs. n=421/738 isolates; median 3 (IQR 2-4) vs. median 2 (1-3) Kruskall-Wallis, p-value<0.001]).

**Fig. 1.**
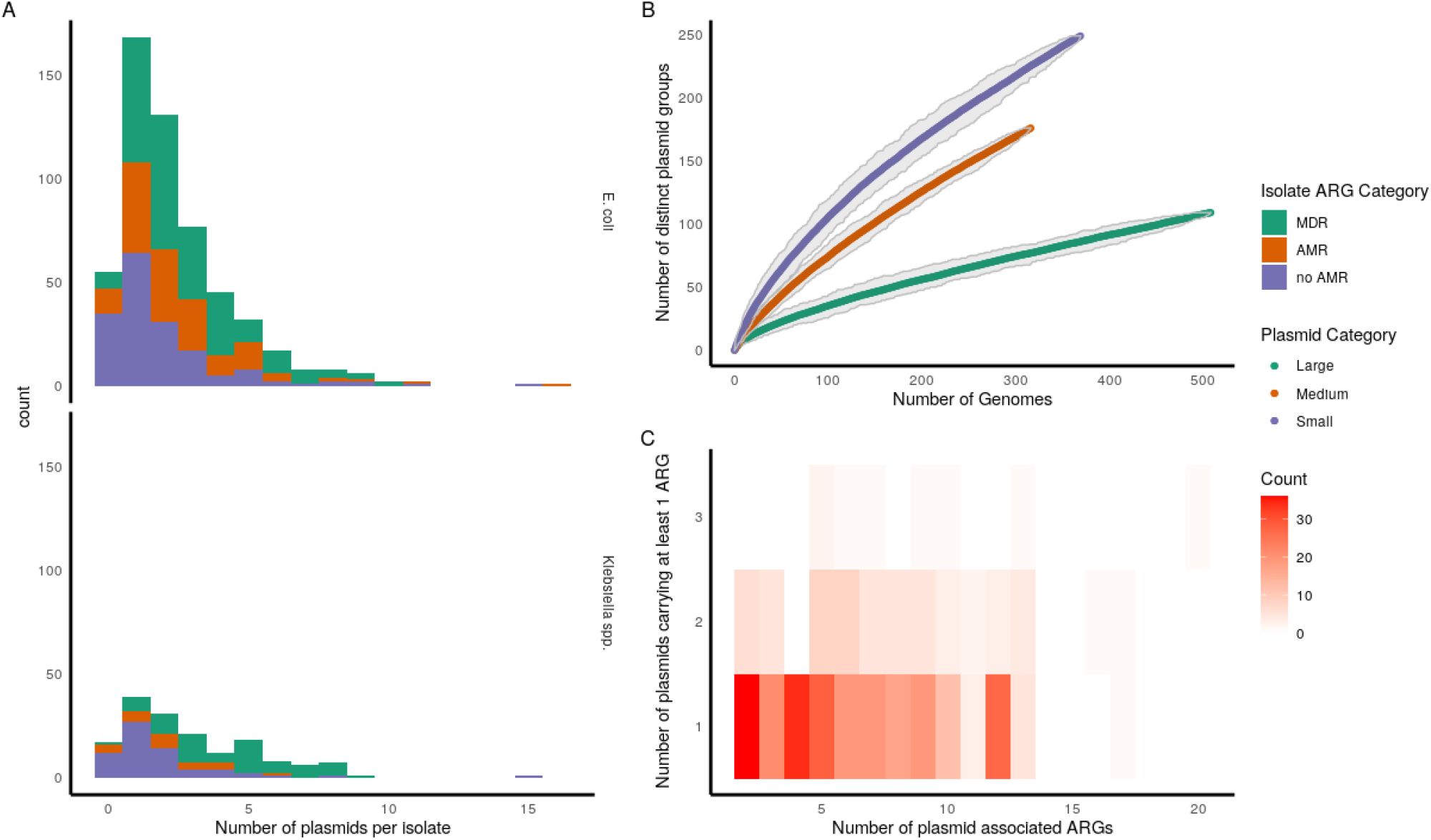
**A** – number of plasmids per isolate for *E. coli* (top panel) and *Klebsiella* spp. (bottum panel), coloured by the number of ARG classes per isolates where MDR is *≥* 3 and AMR 1-2. **B** - Rarefaction curve of the number of novel plasmid groups per new plasmid sequenced stratified by size (Large *≥* 100,000bp, Medium *≥* 10,000 - *<*100,000bp, Small *<* 10,000bp. **C** – Number of plasmid-associated ARGs per isolate vs number of plasmids carrying at least one ARG. Isolates with only one plasmid associated ARG (by definition carried on one plasmid) are excluded.

Despite comprising a relatively small proportion of the total genome (median=2.79%, IQR=1.97-3.97%), plasmids carried 39% (2069/5311) ARGs, 12% (987/8315) virulence genes and 60% (2836/4735) stress response genes. 50% (368/738) isolates carried at least one plasmid-borne ARG and 306 at least 2; of these, 79% (242/306) carried all annotated ARGs on a single plasmid (Figure 1C). In isolates with a medium or large plasmid, co-carriage of a small plasmid was significantly more common in isolates harboring plasmid-borne ARGs 58% (210/361) vs. 46% (119/260) without (Fisher test, p-value=0.003).

### Most BSI isolates carry a large (>100,000bp) plasmid from a small number of common plasmid groups

We first attempted to classify plasmids using existing tools; 17% (317/1880) plasmids could not be assigned a replicon type and 33% (622/1880) had no identifiable relaxase-type. Similarly 26% (487/1880) plasmids were not typable using the recently described Plasmid Taxonomic Unit (PTU) scheme(9); 7% (128/1880) were not typable by any method. Subsequently, we therefore opted to use a previously described classification approach, utilizing a graph-based Louvain community detection algorithm(10) (see methods) which has the advantage of not being reliant on reference databases for group assignation and is thus able to classify all plasmids into groups. This approach yielded 513 groups from 1880 plasmids, of which 164 (32%) contained >1 plasmid, but only 33 (6%) contained ≥ 10 plasmids, and most were singletons (349/513 (68%)). 322/553 (58%) *E. coli* isolates carried a plasmid from one of the four most common, predominantly *E. coli*-associated, large (>=100,000bp) plasmid groups (4/6/7/8) in Figure 3) and similarly 76/161 (47%) *Klebsiella* spp. isolates contained a plasmid from one of the three most common, predominantly *Klebsiella* spp.-associated, large plasmid groups (1/2/5) in Figure 3).

### Plasmid groups are structured by host phylogeny but there is evidence of intra and inter-species transfer events

We found strong evidence that the plasmidome of BSI isolates was structured by host phylogeny, although there was also vast and persistent background diversity. Overall, 141/513 (27%) groups were found in >1 MLST and 22/513 (4%) were found in more than one species; multispecies groups had ≥ 10 members significantly more commonly (8/22 (36%) vs 25/491 (5%), p<0.001) (Figure 3). Sequence type and host species explained 8% and 7% (Adonis p=0.001 for both) of the observed plasmidome variance respectively. ARG content explained a comparatively small amount of variance (R^2^=2%, p=0.001) as did year of isolation (0.03%, p=0.005) and source attribution (R2=1.2%, p=0.99, i.e. suspected focus of infection, only available for a small subset of isolates [198/738]) (Figure 2, panels a, b, c and d respectively). When we focussed on plasmid groups found in the most common *E. coli* STs (131/95/73), we observed that most were seen in only a single ST (78/109) but 13 ‘generalist’ groups were seen in all three STs, and accounted for the majority of plasmids (215/400 (54%)). Highly similar plasmidomes were seen in genetically divergent members of each ST, consistent with multiple horizontal transfer events (Figure S2). Persistent plasmid groups seen in both 2009 and 2018 were also seen in more phylogenetically diverse isolates within STs (Figure S3), consistent with the hypothesis that the persistence of plasmids is linked to their host range potential.

**Fig. 2.**
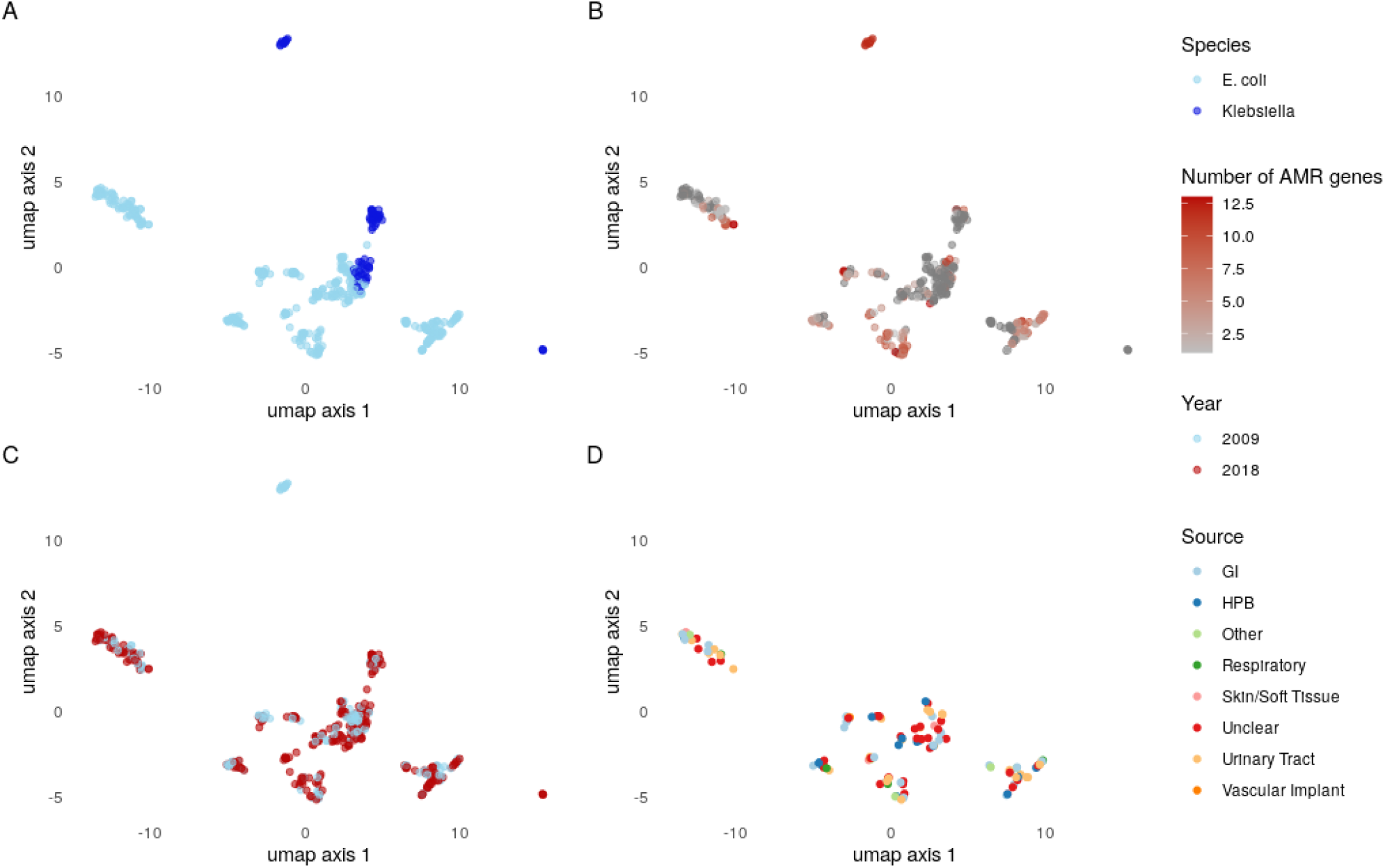
A Umap projection of the mash distances between the plasmidomes of isolates (each point represents the plasmidome, i.e. all plasmid sequences of a single isolate). These are coloured to show the variability explained by species **(A)**/ARG carriage **(B)**/year **(C)** and infection source **(D)**.

**Fig. 3.**
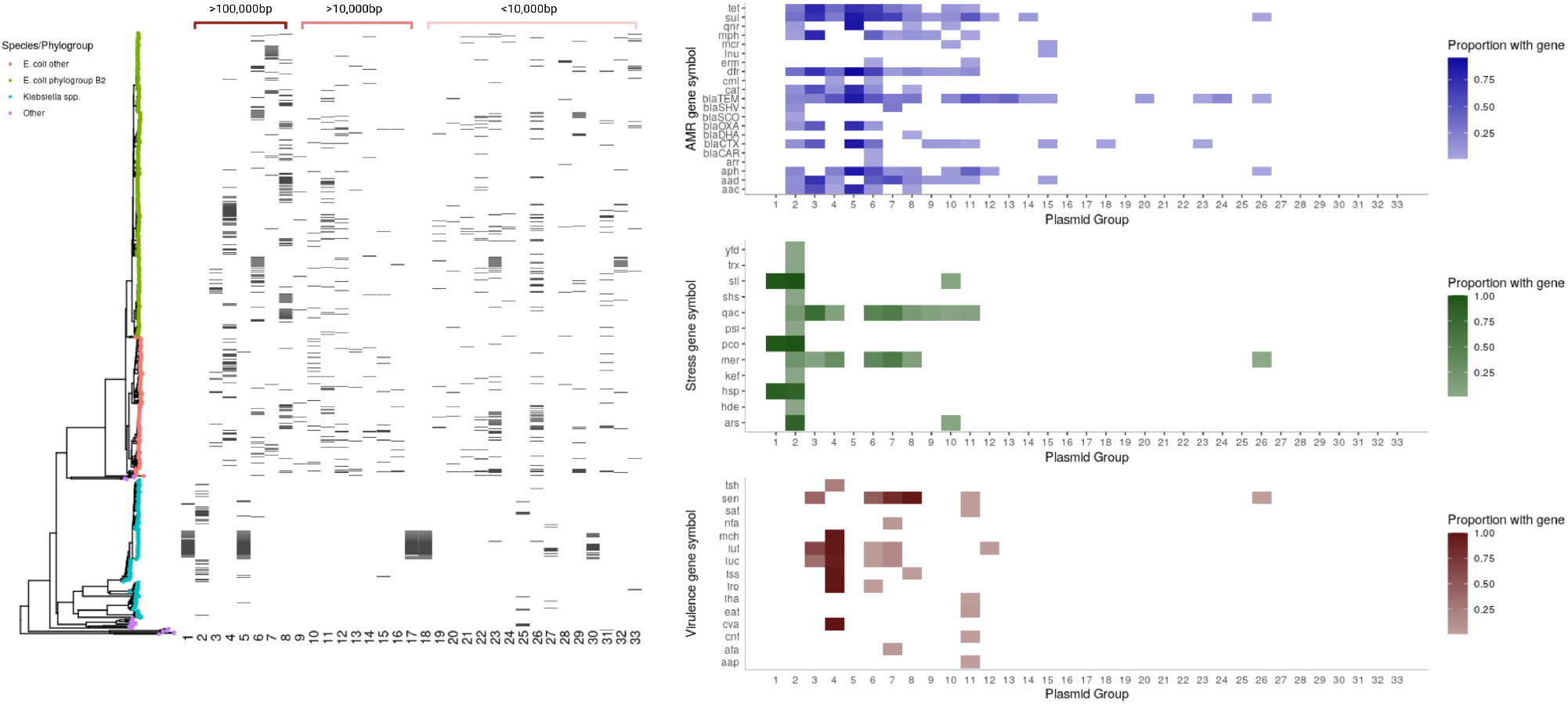
Phylogenetic distribution of the most common (n>=10 members) plasmid groups (n=33 groups) and the content of these. The tree is a neighbour joining tree built on Mash distances between chromosomes. Tip colours represent species/phylogroup. The black bars represent the presence or absence of plasmid groups (shown along the bottom x-axis) for each isolate in the tree. The right panel shows the percentage of isolates within each of these 33 plasmid groups carrying the genes indicated (darker colours denote higher proportion of isolates carrying gene). To improve readability, gene groups have been clustered together.

### Common plasmid groups share genes with each other; gene sharing with chromosomes is also frequent

Whilst we observed only 4% (22/5143) plasmid groups were shared between species, we hypothesised that this might greatly under-represent the true extent of plasmid-mediated horizontal gene transfer given the role of smaller mobile genetic elements and the fact that BSIs represent a tiny fraction of the overall ecological landscape. We therefore looked for evidence of overlap in the pangenome between different plasmid groups as well as between these and host chromosomes. Most genes in the pangenomes of common (i.e. containing n ≥ 10 plasmids) plasmid groups of *E. coli* and *Klebsiella* spp. were non-unique to their group (median % non-unique genes 88%, IQR 67-98%). Most overlap occurred amongst genes found in the plasmid pangenome from the same species (median % shared genes 86% (IQR 50-95%) vs 31% (8-43%) from different species, p<0.001). There was also substantial overlap between plasmid group pangenomes and the chromosome pangenome, although there was some evidence of convergence in the chromosomally integrated mobilome between species, evidenced by less difference in the proportion of genes shared with the chromosome for same vs different species (Figure S4, median 33% (IQR 0-45%) vs 21% (0-35%), respectively p=0.34).

### Plasmids associated with ARG carriage are often highly similar to those with no such genes

The 439 plasmids carrying at least one ARG were predominantly large (≥ 100,000bp, 277/439, 63%), low copy number (median 1.80 IQR 1.63-2.37) and conjugative (347/439, 79%). Whilst most plasmid-borne ARGs were carried by plasmids clustering in a small number of groups (i.e. 81% 1674/2069 ARGs were carried by 8 plasmid groups), 36% (170/474) plasmids in these groups did not carry an ARG and all groups had at least one such member, highlighting that acquisition of ARGs in ARG-negative plasmid backbones represents a common risk across genetically divergent plasmid groups (Figure 4). We repeated this analysis using group assignations given by COPLA (Plasmid Taxonomic Units) and Plasmidfinder (replicon typing) and found similar results (Table S1), suggesting that this finding is robust to the choice of clustering method.

**Fig. 4.**
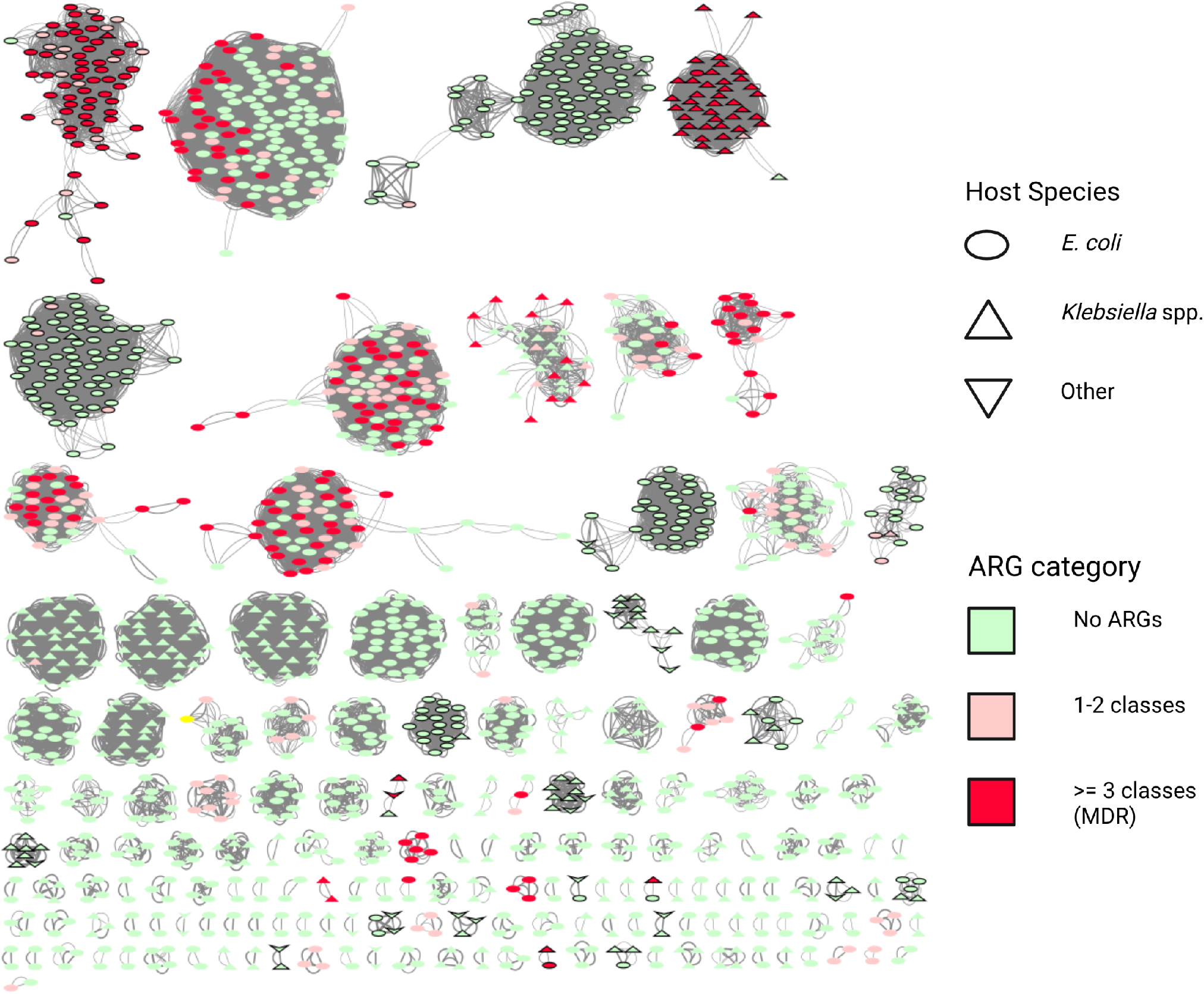
Plasmid network where plasmids (nodes) are connected by edges if they cluster in the same group using the Louvain-based methodology and coloured acording to the number of classes of ARGs that they carry. Edge thickness is drawn proportional to the Jaccard distance (see methods) between plasmids. Multi-species clusters are donated by black outlined shapes. Only plasmids groups with *≥* 2 members are shown.

### Hybrid assembly reveals complex nested diversity associated with key AMR genes, significant chromosomal integration of ARGs and presence of multiple copies in different contexts

Chromosomal integration of ARGs was common: for example, in *E. coli*, 56% (23/41) *bla*_*CTX-M-15*_, 9% (2/22) *bla*_*CTX-M-27*_, 14% (42/293) *bla*_*TEM-1*_, 42% (14/33) *bla*_*OXA-1*_, 39% (7/18) *aac(3)-IIa* and 5% (3/65) *dfrA17* were chromosomally integrated. There was significantly more chromosomal integration of ARGs also seen at least once in a plasmid in our study in *E. coli* vs *Klebsiella* spp. (restricting to 2009 and 2018 only 15% [324/2103] vs 8% 39/478 [8%], Chi-squared test p<0.001). For *E. coli*, there was significantly more chromosomal integration in 2018 vs 2009 (19% [285/1485] vs 6% [39/618], Chi-squared test p<0.001) but there was no evidence of this for *Klebsiella* spp. (7% [13/190] vs 6% [17/279], Chi-squared test p=0.89). For most of these ARGs, there were multiple instances of isolates carrying two (and occasionally more) copies (9 such examples for *bla*_*CTX-M-15*_ (Figure 5), 1 *bla*_*CTX-M-27*_, 29 *bla*_*TEM-1*_, 2 *aac(3)-IIe* and 1 *dfrA7*).

**Fig. 5.**
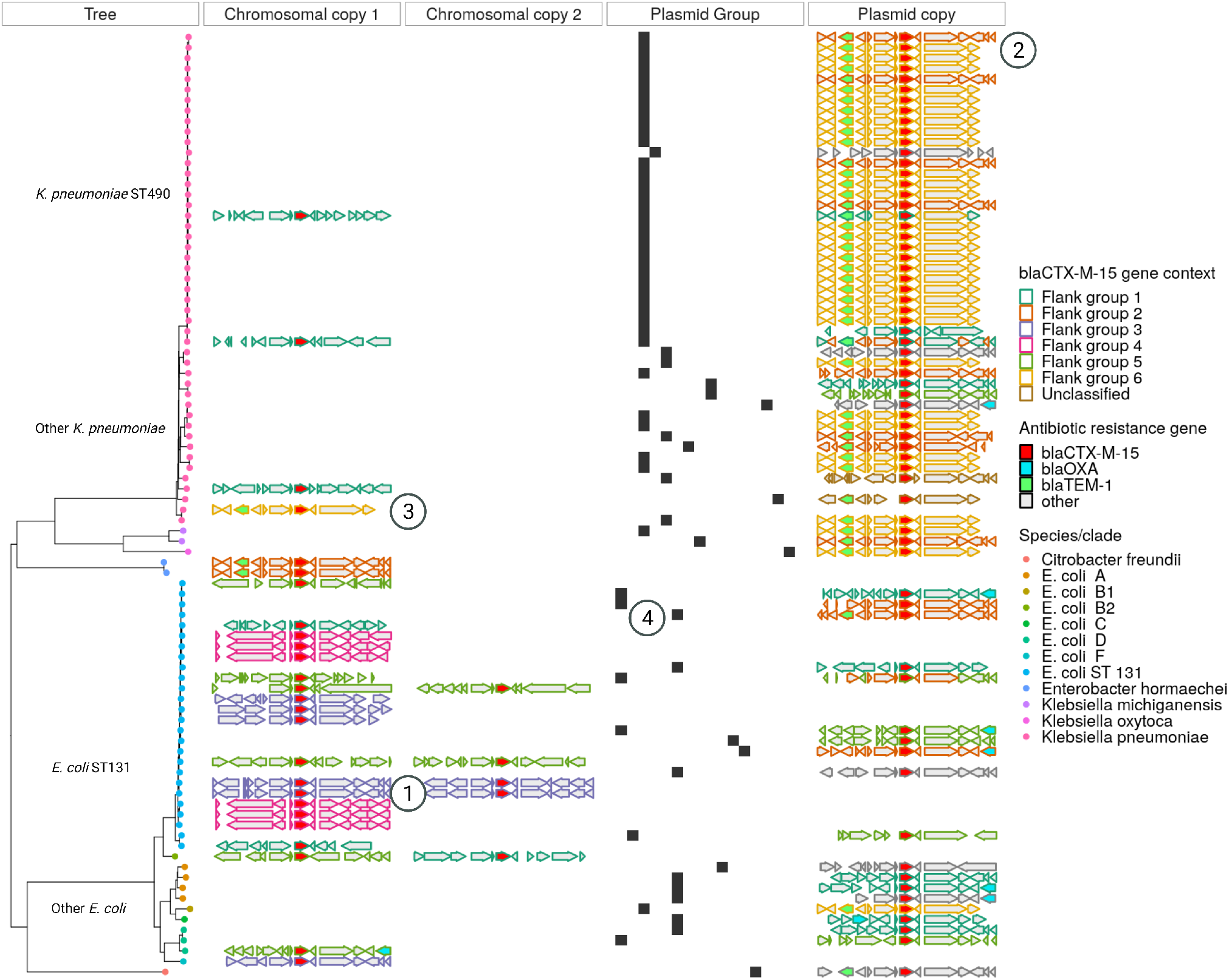
Nested genetic complexity associated with *bla*_*CTX-M-15*_ mobilisation. The ‘Tree’ panel shows a neighbour-joining tree of Mash distances between chromosomes for isolates carrying a *bla*_*CTX-M-15*_ gene. Tip colours represent species/ST/phylogroup. The chromosomal copy 1 and 2 panels show the genetic context 5000bp up- and downstream from chromosomal copies of the *bla*_*CTX-M-15*_ gene (shown in red); the plasmid copy panel shows this equivalent information for isolates carrying a plasmid-borne copy of this gene. The outlining colour in these panels shows the hierarchical cluster assignment of these flanking groups. The plasmid group panel shows group membership of plasmids carrying the *bla*_*CTX-M-15*_ gene with each x-axis position representing a distinct group and black bars showing the presence or absence of these for isolates in the tree. The encircled numbers denote: 1 - different flanking sequences in the same ST, 2 - different flanking sequences in the same plasmid group, 3 - the same flanking group found in both chromosomal and plasmid contexts and 4 - different plasmid groups harbouring the gene found within the same ST.

Given the global importance of the ESBL gene *bla*_*CTX-M-15*_ conferring third generation cephalosporin resistance, we focused on its genetic background and putative dissemination mechanisms. As above, plasmid groups carrying this gene in our dataset were generally species constrained. However, within a single species, considering phylogroup, sequence type and even plasmid group, *bla*_*CTX-M-15*_ was found in a variety of genetic contexts (Figure 5). For example, in *E. coli* ST131 it was found in five plasmid groups and was chromosomally intergrated in 41% (17/41) isolates. Within ST131 sub-clades, there was some evidence of vertical transmission, as well as numerous independent integration events. In many cases, several unique gene flanking regions were found in association with *bla*_*CTX-M-15*_ within a single plasmid group, or identical flanking regions were shared across plasmid groups and between plasmid groups and chromosomes. Visual inspection of gene flanking regions and hierarchical clustering of a weighted graph (Methods: Bioinformatics) suggested that whilst there was substantial diversity, these flanking regions appear to have evolved in a stepwise manner with bilateral association of *bla*_*CTX-M-15*_ and Tn2 in flanking groups 2, 3 and 6 compared to the presence of Kpn14 (groups 1 and 5) and IS26 (group 4) (Figure S5). Inspection of core-genome phylogenies of the two largest *bla*_*CTX-M-15*_ carrying plasmid groups (plasmid groups 2 and 3 in Figure 3) demonstrated multiple probable independent horizontal acquisition events of this gene (and other ARG cassettes Figure S6 and Figure S7), suggesting that a flexible capacity to acquire ARGs through diverse mobile genetics elements rather than a fixed association with them might be important factors for the successful dissemination of the host plasmid.

### Comparison with wider plasmid datasets highlights undersampled plasmid diversity, more widespread inter-species and inter-niche plasmid sharing, and the potential for carbapenemase dissemination amongst “high-risk” plasmid groups

We repeated our graph-based plasmid clustering method on a combined dataset of Oxfordshire plasmids (N=1880, hereby referred to as the ‘Oxfordshire dataset’) and the Global collection of plasmids deposited in the NCBI (N=10,159, denoted the ‘global dataset’) using the same sparsifying threshold (≤ 0.551). This yielded 5913 groups of which 484 contained at least one plasmid from the ‘Oxfordshire dataset’; of these, 326 groups (67%) containing 536 plasmids appeared to be unique to Oxfordshire. 79/484 (16%) of groups containing Oxfordshire plasmids were found in more than one species in the full dataset; of these 57 (72%) occurred in only a single species in the Oxfordshire dataset, highlighting the substantial underestimation of wider between-species dissemination by investigating only a single region and single source (i.e. bloodstream infections).

A striking feature of the global network was that plasmids carrying carbapenemase genes clustered with those that did not (Figure 6). Of 122 plasmid groups with at least one member carrying a carbapenemase gene, 19 (16%) contained at least one Oxfordshire plasmid. These included representatives from the *K. pneumoniae* MDR-associated Oxfordshire BSI dataset groups 2 and 5 (Figure 3), three large groups (Figure 3 groups 3/6/8) widely distributed amongst *E. coli* isolates and two groups of smaller plasmids (<100,000bp, Figure 3 groups 10 and 12), also widely distributed in Oxfordshire *E. coli*. Although only 2% (7/414) Oxfordshire plasmids falling into these groups actually carried a carbapenemase ARG, this suggests the potential for carbapenemase acquisition and dissemination amongst widespread “high-risk” plasmid backbones.

**Fig. 6.**
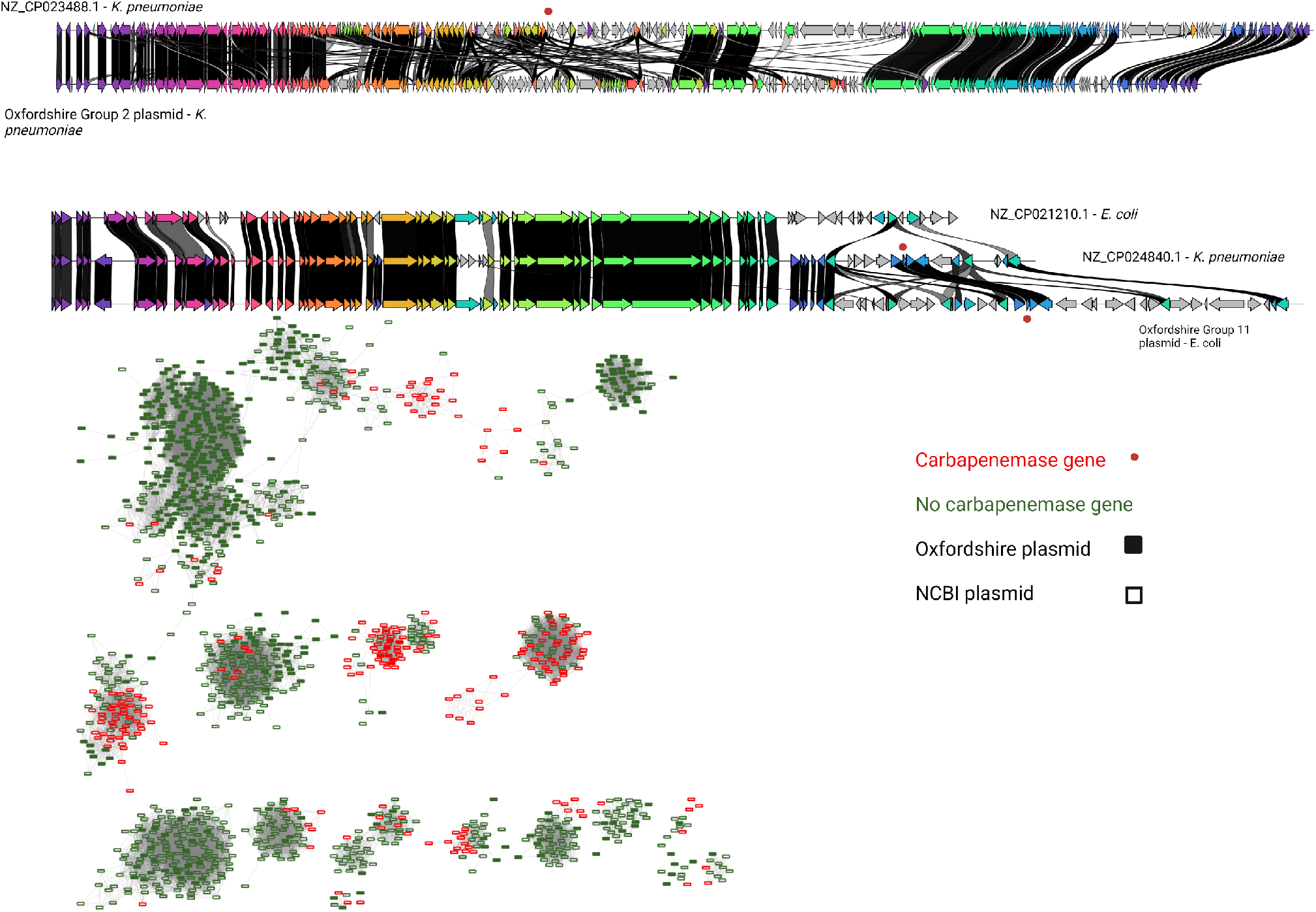
Plasmids carrying carbapenemase genes are highly similar to plasmids without these genes found in Oxfordshire BSIs. A) Each horizontal bar represents a plasmid assembly either from Oxfordshire ‘Group 2/11’ or the NCBI global dataset. Common genes are shown in color (with blast identity between these shown from light grey to black (where the latter represents a perfect match) whereas genes unique to a given plasmid are shown in grey. B) a network plot of plasmids which cluster with carbapenemase carrying plasmids in the global network analysis. Plasmids (nodes) are connected with an edge where the edge weight is *≤*0.551 (see methods). The thickness of edges is displayed so that it is proportional to the edge weight.

### Factors predictive of plasmid group “success”

Having demonstrated that most isolates carry a plasmid from a relatively small number of plasmid groups, we next sought to understand what factors might be driving the widespread dissemination of these amongst BSI isolates. Multivariable Poisson regression analysis revealed that plasmid group frequency (a subjective marker of evolutionary “success”) was associated with isolation in multiple species (adjusted rate ratio aRR 4.89, 95%CI 4.29-5.57, p<0.001), capacity to conjugate (aRR 1.73, 95%CI 1.47-2.04) or mobilise (aRR 1.29, 95%CI 1.13-1.48) (i.e. containing either a relaxase or *oriT* but missing a mate-pair formation marker), carriage of multiple ARGs (aRR 1.23, 95% CI 1.19-1.27)/virulence (aRR 1.44, 95%CI 1.36-1.53)/Toxin-antitoxin genes (aRR 1.32, 95%CI 1.18-1.47) and a higher GC content (aRR 1.01, 95%CI 1.00-1.03) (Table S2). Carriage of ARGs (adjusted Odds Ratio, (aOR=2.88, 95%CI 1.53–5.41, p<0.001) and isolation in multiple species (aOR=7.79, 95%CI 3.07-22.90, p<0.001) were independently associated with a higher probability of plasmid groups being observed internationally (Table S3).

### Machine learning allows risk stratification of plasmids

Given that we have shown that plasmids carrying ARGs are often very similar to those with no such genes (“ARG-negative plasmids”), we hypothesised that it might be possible to predict whether ARG-negative plasmids pose a risk for eventual association with ARGs. To do this we first performed a genome-wide association study (corrected for population structure and plasmid size, using the Oxfordshire dataset only) to identify genes (excluding known ARGs) significantly more or less likely to be carried by plasmids in ARG-associated groups (i.e. plasmid groups where at least one member carries at least one ARG). This revealed significant associations between ARG-associated plasmid groups and presence of insertion/transposon sequences, various virulence factors, toxin/anti-toxin system and heavy metal resistance genes (Table S4).

We then tested the predictive value of these elements to identify ARG-negative plasmids belonging to ARG-associated groups using a variety of models on the Oxfordshire dataset (see methods) with stratified 10-fold cross-validation to estimate out of sample performance. The best performing model (Random Forrest) had a mean accuracy of 90.3% (standard deviation [SD] 2.4%), mean area under the receiver operator curve [AUC] 0.90 (SD 0.02), mean sensitivity 86% (SD 4.3%) and mean specificity 93.4% (SD 2.9%). We re-trained the Random Forrest model on ARG-negative plasmids in the global dataset using only plasmids sequenced prior to 2018 and subsequently made predictions on the held-out 2018 plasmids. This demonstrated that the model generalised well but was less sensitive on this dataset (accuracy 84.6%, AUC 0.82, sensitivity 73.7% and specificity 89.9%).

## Discussion

In this study, we fully reconstructed 738 isolates (1880 plasmids) to conduct the largest, most unselected and comprehensive evaluation of the epidemiology and function of plasmids associated with Gram-negative isolates causing blood-stream infections to date. Most isolates in this study carried a large plasmid from a small number of plasmid groups; these were frequently, but not invariably, associated with carriage of multiple antibiotic resistance, virulence and heavy metal resistance genes, potentially providing survival and fitness benefits to the host bacterium. The fact that most isolates with multiple plasmid-borne ARGs (often from several different classes) carry all of these on a single plasmid reinforces the importance of good antimicrobial stewardship and avoiding unnecessary exposure to all classes of antibiotics to control co-selection as much as possible. Crucially we also found that plasmids carrying ARGs frequently cluster in large, widely disseminated groups with plasmids without these genes, representing a potential set of “high-risk” backbones for ARG acquisition and horizontal spread.

To date, most similar sequencing studies have focused on plasmids carrying particular ARGs (particularly those with ESBL/carbapenem resistance genes) and have therefore not considered how these might be related to plasmids without such genes. We hypothesise that plasmid adaptation to coexist with successful lineages often occurs prior to the acquisition of high-risk ARGs, presenting a potential window of opportunity for intervention which is lost if one is solely focused on the presence of these genes. Our data and should therefore motivate a shift away from studies focusing on a single phenotype or gene of interest and towards efforts to identify and track high-risk plasmid groups or other smaller mobile genetic elements and clearly illustrate that such a new surveillance framework must incorporate unselected sampling frames, i.e. not only selecting isolates with particular AMR phenotypes for sequencing. Notably we found an association of small plasmids and medium/large ARG-associated plasmids, suggesting that they may play an important helper role in ARG plasmid persistence/spread, and a more detailed understanding of this possible synergy could be valuable(11).

Whilst plasmid populations were structured, and plasmid groups where mostly constrained to a single species and in some cases species lineages in Oxfordshire, there was also clear evidence of exchange between lineages of a species and different species. Enterobacterales are widely distributed as commensals and in multiple environmental sources; our study sample is thus extremely sparse relative to the whole ecology. Even where we found no evidence that certain plasmids were shared between species in Oxfordshire, our data demonstrated widespread sharing of the plasmid gene repertoire (including ARGs and their flanking regions) with plasmids and chromosomes in other species. The unexpectedly high proportion of isolates with chromosomally integrated ARGs (and apparent increase across the study period for *E. coli*) may either represent a success of plasmids in conferring survival benefits to their host while lowering their own associated fitness cost or a success of the host by lowering its dependence on the presence of the plasmid.

A limitation of this study is that it is from a single region, mitigated in part by comparisons with publicly available datasets. The lower sensitivity for predicting ARG-group association in the global dataset likely reflects its inherent bias and heterogeneity compared to the Oxfordshire dataset as well as the existence of such ARG-associated groups and genes not observed in our setting. The inability to sequence and/or assemble all plasmids from the selected cohort is an additional limitation. Sequencing only bloodstream infection isolates may lead to underestimation of how much sharing of plasmids between species truly occurs given that this represents a highly selected subset of isolates causing severe disease. This is supported by our analysis of the large publicly available dataset, which demonstrated that several groups found only in a single species in our study have previously been seen in other species. Our results also highlight the substantial limitations of previous studies using reference database-based approaches for plasmid typing and demonstrate that fully reconstructed genomes (i.e. long read sequencing data) are essential in order to provide meaningful insight.

In conclusion our study provides the first high-resolution description of the plasmidome associated with *E. coli*/*Klebsiella* spp. bloodstream infections and demonstrates that using long-read data and unselected sampling frames is essential in order to fully appreciate its complexity. Previous studies of plasmid epidemiology in Gram-negatives have primarily focused on MDR/carbapenemase-carrying isolates; our finding that non-ARG carrying plasmids are often highly similar to plasmids isolated in these earlier studies demonstrates the potential for rapid dissemination of ARGs to settings where they are currently rare. We recommend that surveillance is based on unselected sampling frames, long-read sequencing and considers plasmids and smaller mobile genetic elements to develop a representative understanding of the horizontal gene transfer landscape to facilitate appropriate intervention.

## Methods

### Isolate selection

We have previously reported analyses of short read sequencing data from *E. coli* and *Klebsiella* spp. bloodstream infection isolates in Oxfordshire between 2009 and 2018 as described previously(12, 13). In this study, we additionally sequenced all *E. coli* and *Klebsiella* spp. isolates from 2009 and 2018 using Oxford Nanopore Technologies. We also sequenced a subset of isolates from intervening years, using stratified random sampling based on analysis of short-read data to capture maximum plasmid diversity. Additionally we selected isolates from clinically important local AMR-associated outbreaks and representatives of other species with apparently similar plasmidomes. Details of successfully sequenced isolates, those excluded and the stratification and selection methods are available in the appendix.

### Sequencing

DNA for long-read sequencing was extracted either using Qiagen Genomic Tip/100G according to the manufacturer’s instructions, or with the BioMerieux Easymag using the manufacturer’s generic short protocol with a final elution volume of 50 µL. The Qubit 2.0 Flourometer was used to quantify DNA. Sequencing libraries were prepared using the Oxford Nanopore Technologies Native (n=23) and Rapid (all other) barcoding kits, according to the manufacturer’s instructions. Sequencing was performed on GridIons with R9.4 flowells, which were reused multiple times utilising the ONT Flow Cell Wash kit and our previously validated protocol(14). Illumina (short-read) data was created as previously described(13).

### Bioinformatics

Reads were first base-called and demultiplexed using Guppy (v3.1.5, Oxford Nanopore Technologies) with Deepbinner(15) (v0.2.0) subsequently used to recover additional unclassified reads as previously described(14). Our strategy for hybrid assembly is depicted in Figure S8. We first assembled all isolates using Unicycler(15) (–mode bold) with the raw Illumina and ONT reads as input. In parallel we performed another assembly where Unicycler was given an assembly graph from Flye(16) (run with –plasmids –meta and reads which had been polished using Ratatosk(17)) and short reads pre-processed by Shovill(18). The most contiguous assembly of these was used (or the latter if both were complete). If neither hybrid assembly completed then we used the Flye assembly (with four subsequent rounds of Pilon(19) polishing) if this was complete. Incomplete assemblies (where ≥ 1 replicon [i.e. either plasmids or the chromosome] had >1 contig) were excluded from further analysis (n=215).

Rarefaction analysis with performed using the R library Micropan(20). Annotation of genes was performed using AMRFinder Plus(21), ABRicate(22), TADB 2.0(23) and Prokka(24); custom/manually augmented databases (available at www.github.com/samlipworth/GN_BSI_Hybrid) were used for the latter two to attempt to improve the proportion of annotatable toxin-antitoxin systems/plasmid associated genes respectively. GC content and predicted mobility were extracted from Mobsuite output. GC-gap was defined as *GC content of plasmid – GC content of chromosome*.

Pangenomes were analysed using Panaroo(25) (v1.2.8 – clean-mode sensitive) and visualised with a Umap projection created using the R package Umap(26). Variance in the pangenome explained by e.g. AMR content/year/species was examined using a permanova perfomed in the R package vegan(27). Gene flanking regions were analysed using Flanker (v1.0, –w 0 –wstop 5000 –wstep 100)(28). The Reder package(29) was used to cluster a weighted graph created from a matrix in which distances were determined to be the greatest distance from the gene (in both upstream and downstream directions) in pairs of isolates which were in the same Flanker cluster; this analysis was repeated in an all vs all fashion for all isolates. Flanking regions were annotated using the Galileo AMR software (Arc Bio, Cambridge MA USA).

### Plasmid clustering

Robust taxonomic classification of plasmids remains a challenge(30). We therefore used two established methods that have been applied to large-scale short read sequencing datasets, Replicon typing using PlasmidFinder(31) and Relaxase typing with MOB-suite(32). We also typed all plasmids using the recently described Plasmid Taxonomic Unit nomeclature(8) (using COPLA(9)). As a substantial number of plasmids remained unclassified by all these methods, we additionally utilized a recently described graph-based classification system(10). Mash(v2.3)(33) (-s 1000, -k 21) was used to create an all vs. all distance matrix of plasmid assemblies where the distance was taken to be 1 – the proportion of shared kmers between the plasmid of interest and plasmids in the sketch sequences, where plasmids with a distance of 0 share all kmers in the sketch space whereas those with a score of 1 share no common kmers. This was used to create a weighted graph using the R package Igraph(34) where vertices represent plasmids and edges between these are weighted by the distance described above. Community detection on this graph was performed using the Louvain algorithm which seeks to maximise the density of edges within vs between communities. We optimised performance of this algorithm as described previously(10) by sparsifying the graph, removing edges with a weight ≤ a threshold which was selected by iteration. This approach performed optimally (i.e. assigned the maximum number of isolates to larger [n ≥ 10 isolate] clusters) when the graph was sparsified at an edge weight of ≤ 0.551 prior to community detection (Figure S9); this parameter was used for all subsequent analysis. The final sparsification threshold was selected to optimise the number of plasmids assigned to large (n ≥ 10) clusters. We compared the classifications given by this approach to other methods using the Normalized Mutual Information index in the R package NMI(35), which demonstrated good agreement with previously described classification methods using normalised mutual information (NMI; see Methods): replicon-typing NMI=0.81, relaxase-typing NMI=0.93, plasmid taxonomic unit (PTU) NMI=0.81.

### Comparison with existing plasmid sequencing data

To place our plasmid sequencing data in a global context, we downloaded all available plasmids (n=10,159) from a recently curated plasmid collection(7) for comparison. We refer to the Oxfordshire isolates as the “Oxfordshire dataset” and the combined collection as the “Global dataset”. We computed a pairwise distance matrix and performed Louvain-based clustering as described above, sparsifying the graph using the same threshold (0.551) as in the main analysis.

### Statistical analysis

To investigate factors associated with geographical dissemination of plasmid groups we subsetted the global dataset to include only Oxfordshire isolates and those from NCBI not from the UK and where the location of isolation was known. We further filtered this to include only plasmid clusters observed at least once in Oxfordshire. Isolation in more than one country was used as the binary dependent variable in a logistic regression with other plasmid group features (e.g. ARG/virulence/GC content) as independent variables. Multivariable associations between all available plasmid group metrics (independent variables) and plasmid group frequency in the dataset (dependent variable) were estimated using Poisson regression in exploratory analyses. Comparisons of continuous variables and proportions between groups used Kruskal-Wallis/Wilcoxon Rank-sum and Fisher/Chi-squared tests respectively in R version 4.1(36).

To search for non-AMR plasmid-borne genes associated with carriage of ARGs, we performed logistic regression with membership of an ARG-associated group as the dependent variable and each gene in the plasmid pangenome as the independent variable, adjusting for population structure using multi-dimensional scaling (MDS) of mash distances (R package CMD scale), represented in 10 dimensions(37). We additionally adjusted for plasmid size using three categories (’large’ ≥ 100,000bp, medium ≥ 10,000-<100,000bp and small <10,000bp). P-values were adjusted for multiple comparisons using the Bonferroni method after removing genes with <1% population frequency. This pangenome-wide association study was performed using only the Oxfordshire plasmid dataset.

We then tested the predictive value of these genes (n=178) to identify plasmids not carrying ARGs (ARG-negative plasmids) which were found in ARG-associated groups in the Oxfordshire dataset (N=1447 plasmids of which 609 where in ARG-associated groups). We evaluated the performance of nine models (logistic regression, linear discriminant analysis, K neighbours classifer, decision tree classifier, gaussian naive Bayes, random forest classifier and gradient boosting classifier and a voting classifier combining all of these) using 10-fold cross validation, which was repeated 100 times. We further evaluated the performance of the best performing model (random forest) using ARG-negative *E. coli* and *Klebsiella* spp plasmids in the global dataset with the same features as before (significant gene hits from the pangenome GWAS above). We split the dataset into plasmids collected prior to 2018 (global training set on which the model was re-trained n=656 plasmids of which 221 where in ARG associated groups) and those collected subsequently (held-out global testing set on which final metrics were reported N=306 plasmids of which 99 where in ARG-associated groups). This analysis was performed using the SciKitLearn(38) package in Python version 3.7.7.

## Data Availability

All sequencing data has been deposited in the NCBI under project accession number PRJNA604975.

https://www.ebi.ac.uk/ena/browser/view/PRJNA604975

## Data visualisation

Data were visualised using the ggplot2(39) and gggenes(40) packages in R, Clinker(41), Cytoscape(42) and Biorender (www.biorender.com).

## ACKNOWLEDGEMENTS

This study is supported by the National Institute for Health Research Health Protection Research Unit (NIHR HPRU) in Healthcare Associated Infections and Antimicrobial Resistance at the University of Oxford in partnership with Public Health England (PHE) (NIHR200915). WM is supported by a scholarship from the Medical Research Foundation National PhD Training Programme in Antimicrobial Resistance Research (MRF-145-0004-TPG-AVISO). ASW and TEAP are also supported by the NIHR Oxford Biomedical Research Centre. ASW is an NIHR Senior Investigator. NS is an NIHR Oxford BRC Senior Fellow. The views expressed are those of the authors and not necessarily those of the National Health Service, NIHR, Department of Health, or PHE. SL is supported by an MRC Clinical Research Training Fellowship (MR/T001151/1). L.P.S. is a Sir Henry Wellcome Postdoctoral Fellow funded by Wellcome (Grant 220422/Z/20/Z). Computation used the Oxford Biomedical Research Computing (BMRC) facility, a joint development between the Wellcome Centre for Human Genetics and the Big Data Institute supported by Health Data Research UK and the NIHR Oxford Biomedical Research Centre. The authors gratefully acknowledge the assistance of Anthony Brown in performing DNA extractions.

