## Supplement for "The mobilome associated with Gram-negative bloodstream infections: A large-scale observational hybrid sequencing based study"

**Supplementary Methods:**

**Selection of isolates for long read sequencing:**

We previously sequenced all (90-day deduplicated) E. coli and Klebsiella spp. BSI isolates from 2009-2018(Lipworth et al. 2021). We also have a collection of other Gram negative species from August 2011 (unpublished data) which were also sequentially collected without selection for phenotype and deduplicated to 90-days. As already described we intended to sequence all E. coli and Klebsiella spp. isolates from 2009 and 2018. We then analysed the contigs of all remaining short read assemblies using MLPlasmids(Arredondo-Alonso et al. 2018) to classify then as likely plasmid or chromosomal in origin (using –use-full-khash-sets). All likely plasmid contigs where binned together and sketched using Dashing(Baker and Langmead 2019) (default settings) and a distance matrix subsequently collected. We sparsified this matrix at 0.8 and used the LinkComm(Kalinka and Tomancak 2011) package in R to identify communities. We then selected one representative per ST from the largest (n>=10) clusters. Remaining capacity (limited by resource and time as laboratory work took place during the SARS-COV-2 pandemic) was filled using isolates with similar multi-species plasmidomes and local AMR-associated outbreak clones.

**Table S1a – Counts of plasmids with/without ARGs in the largest PTU groups.**

| **Assigned PTU** | **-** | **PTU-?** | **PTU-B/O/K/Z** | **PTU-FE** | **PTU-FK** | **PTU-I1** | **PTU-X4** | **PTU-Y** |
| --- | --- | --- | --- | --- | --- | --- | --- | --- |
| Non-ARG containing | 439 | 40 | 21 | 174 | 24 | 1 | 14 | 0 |
| ARG-containing | 48 | 27 | 37 | 206 | 58 | 3 | 11 | 4 |

**Table S1b – Counts of plasmids with/without ARGs in the largest PlasmidFinder groups.**

| Assigned Rep-type | **Col156** | **IncFIA** | **IncFIB(AP001918)** | **IncFIC(FII)** | **IncFII** | **IncFII_pKP91** | **IncFII(29)_pUTI89** | **IncFII_pRSB107** |
| --- | --- | --- | --- | --- | --- | --- | --- | --- |
| Non-ARG-Containing | 220 | 12 | 154 | 52 | 16 | 37 | 81 | 14 |
| ARG-containing | 107 | 97 | 206 | 74 | 47 | 48 | 41 | 45 |

**Table S2**: Plasmid factors associated with plasmid group frequency in the dataset.

| **Variable** | **Univariable Rate Ratio (95% CI)** | **P value** | **Multivariable Rate Ratio (95% CI)** | **P value** |
| --- | --- | --- | --- | --- |
| Number of species in which the plasmid group was found in | 6.05 (5.41-6.75) | <0.001 | 4.89 (4.29-5.57) | <0.001 |
| Median number of AMR genes | 1.34 (1.30-1.37) | <0.001 | 1.23 (1.19–1.27) | <0.001 |
| Median number of virulence genes | 1.48 (1.41-1.54) | <0.001 | 1.44 (1.36-1.53) | <0.001 |
| Median number of stress genes | 1.00 (0.98-1.02) | 0.99 | 1.00 (0.98-1.02) | 0.87 |
| Median number of known plasmid replicon sequences | 2.73 (2.36-3.16) | <0.001 | 0.83 (0.69-0.98) | 0.032 |
| Median number of toxin/antitoxin genes | 1.54 (1.45-1.64) | <0.001 | 1.32 (1.18-1.47) | <0.001 |
| Size category small (<10,000bp) | - | - | - | - |
| Size category medium (≥ 10,000bp-99,999bp) | 0.77 (0.68-0.86) | <0.001 | 0.53 (0.45-0.63)  0 | <0.001 |
| Size category large (≥ 100,000bp) | 1.62 (1.46-1.79) | <0.001 | 0.63 (0.50-0.79) | <0.001 |
| GC content | 1.03 (1.02-1.04) | <0.001 | 1.01 (1.00-1.03) | <0.001 |
| Non-mobilisable (reference category) | - | - | - | - |
| Mobilisable | 1.73 (1.53-1.96) | <0.001 | 1.29 (1.13-1.48) | <0.001 |
| Conjugative | 2.35 (2.08-2.66) | <0.001 | 1.73 (1.47-2.04) | <0.001 |

**Table S3**. Features associated with plasmid groups being unique to Oxford (N=326 groups) or also found in the Acman/global plasmid dataset (N=158 groups). There were 5429 plasmid groups only found in the global plasmid dataset. In this analysis plasmids from NCBI were excluded if they were isolated in the UK or if the country of origin was unknown. Statistically significant (p<0.05) variables in the multivariable analysis are highlighted in bold.

| **Variable** | **Odds Ratio (Univariable)** | **Lower confidence interval** | **Upper confidence interval** | **p value** | **Odds Ratio (Multivariable)** | **Lower confidence interval** | **Upper confidence interval** | **p value** |
| --- | --- | --- | --- | --- | --- | --- | --- | --- |
| **Antimicrobial resistance gene present (ARG)** | **3.46** | **1.98** | **6.02** | **<0.001** | **2.88** | **1.53** | **5.41** | **<0.001** |
| Virulence gene | 1.37 | 0.56 | 3.08 | 0.46 | 1.63 | 0.57 | 4.35 | 0.34 |
| Stress response gene | 1.77 | 1.00 | 3.07 | 0.04 | 1.58 | 0.68 | 3.66 | 0.29 |
| Medium Size (10,000-99,999bp) | 0.92 | 0.54 | 1.53 | 0.75 | 0.55 | 0.26 | 1.11 | 0.11 |
| Large Size (≥ 100,000bp) | 1.41 | 0.82 | 2.41 | 0.21 | 0.61 | 0.22 | 1.62 | 0.34 |
| Conjugative | 2.20 | 1.25 | 3.90 | 0.01 | 1.71 | 0.84 | 3.57 | 0.15 |
| Mobilisable | 1.68 | 0.99 | 2.91 | 0.06 | 1.24 | 0.69 | 2.25 | 0.48 |
| GC content | 1.06 | 1.01 | 1.12 | 0.01 | 1.04 | 0.99 | 1.10 | 0.14 |
| **Number of species in which the plasmid was found in the Oxfordshire dataset** | **9.44** | **3.91** | **26.70** | **<0.001** | **7.79** | **3.07** | **22.90** | **<0.001** |

**Table S4**: Genes/gene clusters associated with ARG-carrying plasmids. All values report univariable logistic regression adjusted for plasmid size (categorical large/medium/small) and population structure (10 dimensions of multidimensional scaling of Mash distances between host chromosomes). P values have been adjusted by the Bonferroni method; only significant (p<0.05) results are shown for genes annotated as something other than “hypothetical protein”. Annotations are as given by Prokka/Panaroo. Where Panaroo clustered together multiple prokka annotations, only the first of these is shown. Two plasmids (sizes 1570 and 1308) were excluded from the analysis because they had no coding regions identified. Results are ordered by p value.

| **Gene** | **N in ARG-carrying plasmids** | **N in non-ARG-carrying plasmids** | **OR** | **P value** |
| --- | --- | --- | --- | --- |
| psiB | 437 | 51 | 17.52 | <0.001 |
| iucD | 105 | 8 | 21.57 | <0.001 |
| insB_1 | 327 | 178 | 18.69 | <0.001 |
| shiF | 97 | 8 | 20.95 | <0.001 |
| iucB | 97 | 8 | 20.95 | <0.001 |
| tnpA | 272 | 53 | 6.47 | <0.001 |
| sitD | 96 | 3 | 53.63 | <0.001 |
| sitC | 96 | 3 | 53.63 | <0.001 |
| sitB | 95 | 3 | 53.61 | <0.001 |
| sitA | 93 | 3 | 53.56 | <0.001 |
| ntdC | 83 | 2 | 91.82 | <0.001 |
| orf156_1 | 331 | 22 | 6.28 | <0.001 |
| orf157_1 | 304 | 21 | 6.60 | <0.001 |
| cvaA | 86 | 2 | 91.07 | <0.001 |
| KF37082 | 430 | 53 | 3.99 | <0.001 |
| repA_1 | 269 | 56 | 195.73 | <0.001 |
| areA | 441 | 49 | 8.04 | <0.001 |
| hha | 32 | 20 | 75.45 | <0.001 |
| pemK_2 | 185 | 4 | 33.70 | <0.001 |
| pemI | 185 | 4 | 33.70 | <0.001 |
| orf36 | 34 | 4 | 23.24 | <0.001 |
| yjcC_3 | 32 | 2 | 62.29 | <0.001 |
| ycgB_1 | 452 | 54 | 3.62 | <0.001 |
| arsH | 29 | 4 | 24.13 | <0.001 |
| orf34 | 28 | 4 | 24.11 | <0.001 |
| repI | 342 | 32 | 5.21 | <0.001 |
| yuaT | 92 | 2 | 59.47 | <0.001 |
| orf05 | 358 | 93 | 4.20 | <0.001 |
| parM_1 | 45 | 3 | 47.37 | <0.001 |
| tnpA | 78 | 4 | 21.27 | <0.001 |
| ydfA | 292 | 24 | 4.57 | <0.001 |
| ydeA | 340 | 53 | 3.51 | <0.001 |
| higA_1 | 73 | 13 | 7.79 | <0.001 |
| insE | 18 | 33 | 0.09 | <0.001 |
| yehA | 310 | 26 | 4.84 | <0.001 |
| topIII_1 | 28 | 17 | 32.11 | <0.001 |
| higB | 73 | 12 | 7.73 | <0.001 |
| traE_1 | 28 | 7 | 11.70 | <0.001 |
| tnpA_3 | 113 | 143 | 0.23 | <0.001 |
| etsA | 85 | 11 | 8.88 | 0.0012 |
| etsB | 85 | 11 | 8.88 | 0.0012 |
| etsC | 85 | 11 | 8.88 | 0.0012 |
| mob2 | 17 | 83 | 9.85 | 0.0013 |
| hnh | 10 | 76 | 0.12 | 0.0014 |
| tnpA_3 | 73 | 3 | 38.33 | 0.0018 |
| TF134_00008 | 95 | 1 | 153.05 | 0.002 |
| hlyF | 86 | 12 | 8.03 | 0.002 |
| celG | 3 | 20 | 0.04 | 0.0025 |
| repL | 12 | 8 | 14.98 | 0.0027 |
| ant | 12 | 8 | 14.98 | 0.0027 |
| topB_1 | 32 | 29 | 34.37 | 0.004 |
| ppfA | 13 | 8 | 14.39 | 0.0046 |
| upfB | 13 | 8 | 14.39 | 0.0046 |
| ydcB | 463 | 83 | 2.97 | 0.0056 |
| pdcB | 58 | 10 | 10.48 | 0.0057 |
| pDCA | 58 | 10 | 10.48 | 0.0057 |
| mobA | 72 | 29 | 9.45 | 0.0058 |
| doc | 58 | 11 | 9.53 | 0.008 |
| ybaA | 258 | 13 | 5.05 | 0.0083 |
| arsD | 29 | 12 | 6.75 | 0.0084 |
| arsA_2 | 29 | 12 | 6.75 | 0.0084 |
| cra | 13 | 8 | 13.09 | 0.0091 |
| int | 379 | 113 | 3.97 | 0.0092 |
| int2 | 99 | 25 | 4.29 | 0.01 |
| pmgS | 13 | 8 | 12.52 | 0.013 |
| hdmD | 13 | 8 | 12.52 | 0.013 |
| yacA | 130 | 21 | 4.60 | 0.013 |
| toxin_1 | 9 | 34 | 0.11 | 0.015 |
| traY | 124 | 11 | 5.98 | 0.017 |
| arsA | 30 | 13 | 6.05 | 0.018 |
| traJ | 123 | 11 | 5.96 | 0.018 |
| ydiL | 190 | 10 | 6.77 | 0.024 |
| dnaQ | 91 | 7 | 7.23 | 0.038 |
| mat | 13 | 8 | 11.44 | 0.039 |
| traM_2 | 511 | 114 | 2.94 | 0.042 |

**upplementary Figure Legends**

**Figure_S1:** Breakdown of isolates included/excluded in the study by species.

**Figure_S2**: The similarity of the plasmidome is negatively correlated with phylogenetic distance within an ST. Left panel: plasmid similarity in terms of the presence/absence of plasmid clusters. Right panel: plasmid similarity in terms of the presence/absence of plasmid-borne genes. In both cases, the plasmidome similarity between two isolates is 1 minus the Jaccard index (with the Jaccard index involving isolates carrying no plasmids defined as 1).

**Figure_S3:** For each of the three selected *E. coli* STs, all the plasmid groups seen in isolates of that ST are shown. Each point shows the median phylogenetic distance between isolates carrying that plasmid cluster, subsetted by the number of STs the plasmid cluster was seen in.

**Figure_S4:** Overlap between plasmid and chromosomal pangenomes. Each dot represents a single plasmid group (only the largest [n≥10] plasmid groups are shown). The x-axis denotes the predominant host species in which the plasmid group was isolated and the fill colours denote the comparator species. The y-axis position denotes the proportion of genes in the pangenome (as annotated by Prokka/Panaroo) shared between genetic contexts (i.e. plasmid versus chromosome) as indicated. The colour of the dots shows the median plasmid group size.

**Figure_S5**: Hierachical clustering of a weighted graph formed from a distance matrix where the distance between each pair of plasmids was the maximum window length at which flanker placed them in the same cluster by single linkage. The resulting dendrography was cut at 0.5 and groups (and nested groups) are shown as identified by RedeR. The lowest levels of these have been renamed sequentially (FG – flank group) and colour coded to match the colours used in Figure 4. The annotations as given by Galileo AMR are shown below for each flank group.

**Figure_S6**: Core genome phylogeny and gene presence/absence heatmap (for genes in the AMRFinder/PlasmidFinder databases). For plasmid group 2 (Figure 2).

**Figure_S7**: Core genome phylogeny and gene presence/absence heatmap (for genes in the AMRFinder/PlasmidFinder databases). For plasmid group 3 (Figure 2).

**Figure_S8:** Pictorial representation of assembly pipeline used. Option 1 assemblies were used in preference to option 2, which were preferred to option 3.

**Figure_S9**: Optimisation of threshold and method using to assign plasmids to plasmid groups. The top two rows show the results of a single linkage methodology whereas the bottom two show results obtained using the Louvain clustering algorithm used in our analysis. X-axis shows the Mash distance used to sparsify the group. Number of communities represents the number of plasmid groups n≥10, whereas the largest community represents the largest plasmid group formed at a given threshold. NMI – Normalised mutual information, PTU – plasmid taxonomic unit. The red hashed line shows the Mash distance threshold of 0.551 which was used in the final analysis.
