## Supplementary figures and images for "The mobilome associated with Gram-negative bloodstream infections: A large-scale observational hybrid sequencing based study"

### Fig_S1_new.png

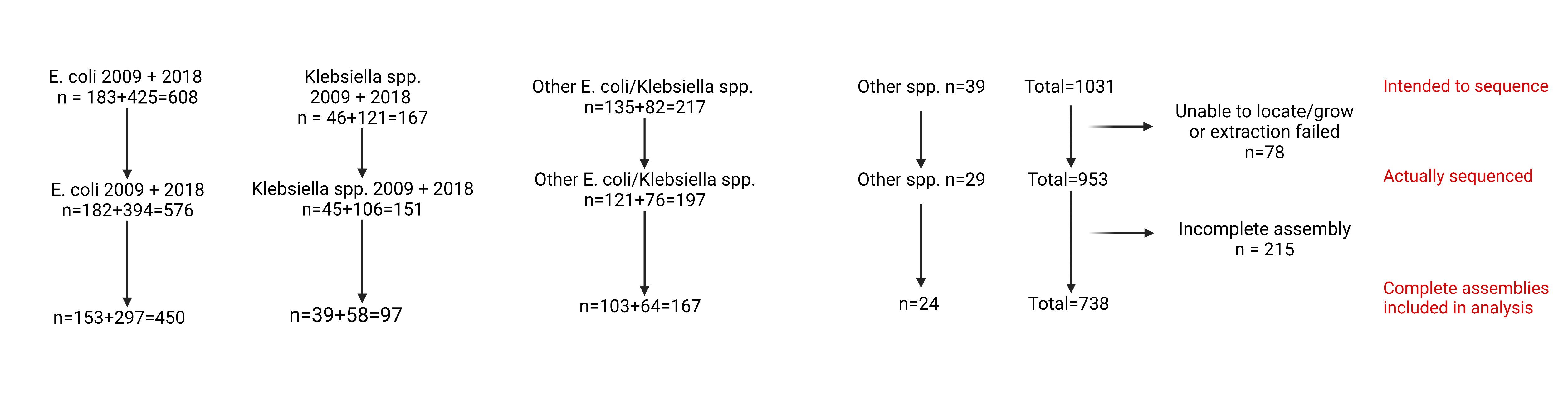

### Fig_S2_new.png

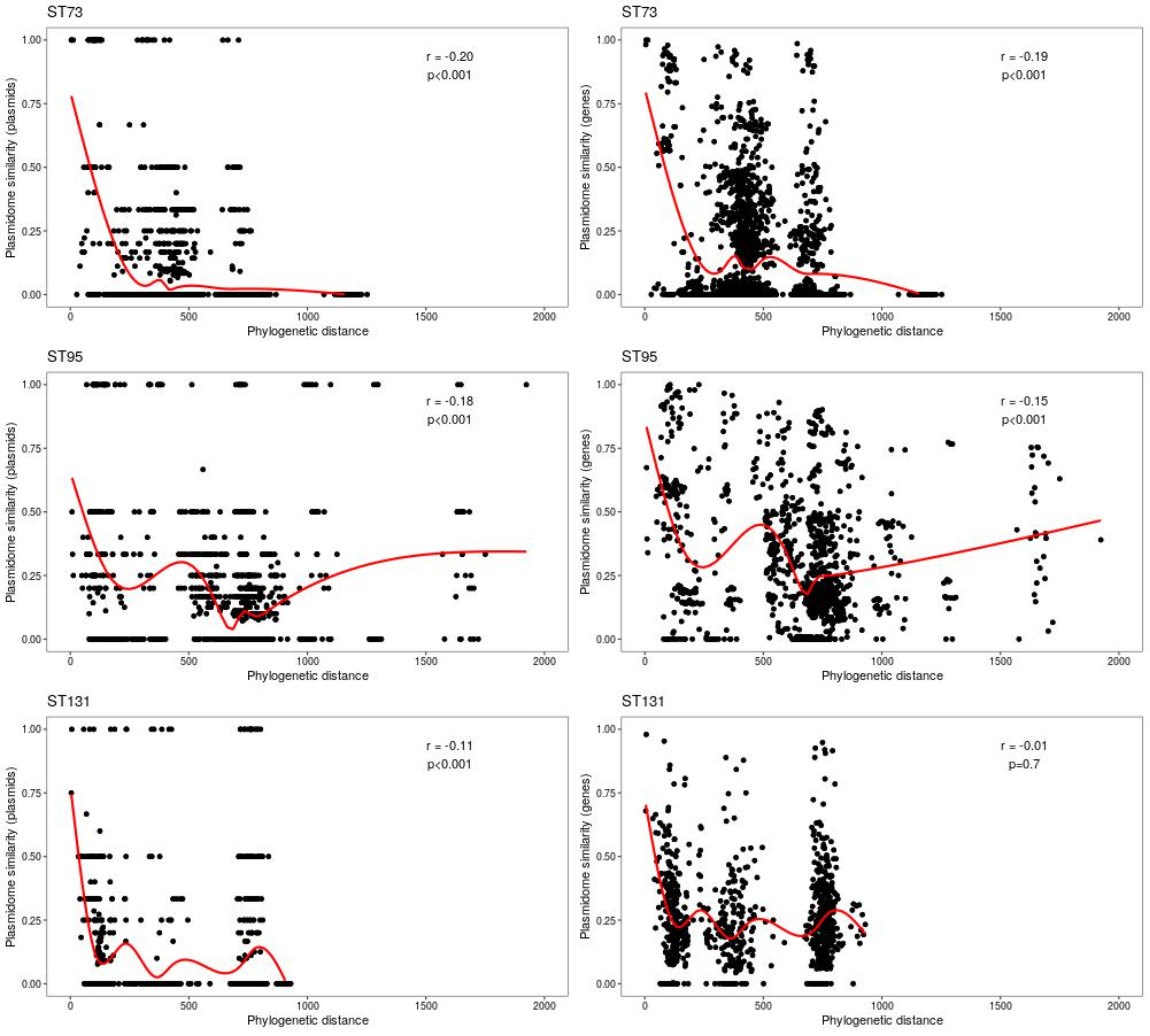

### Fig_S4_new.png

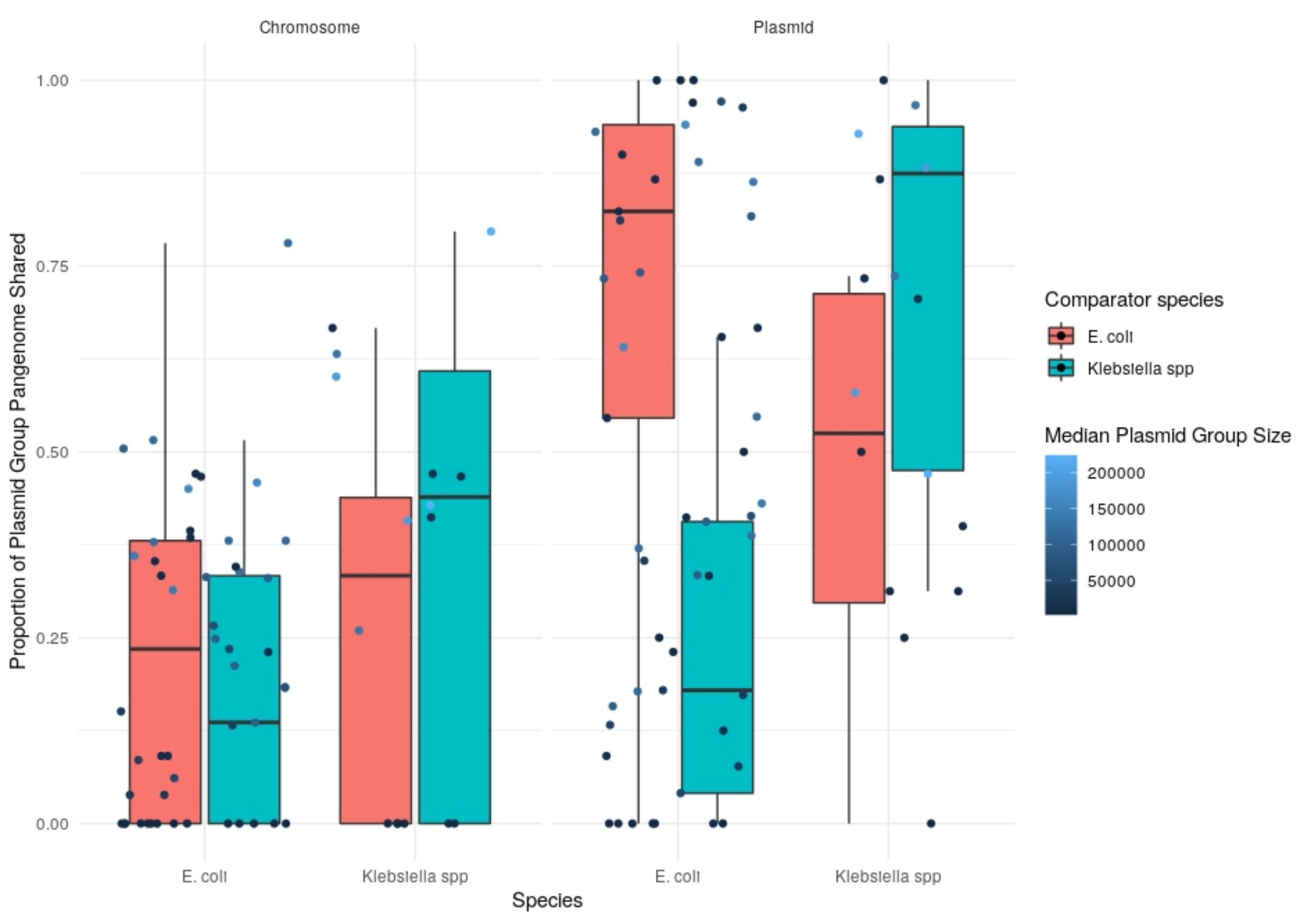

### Fig_S6_new.png

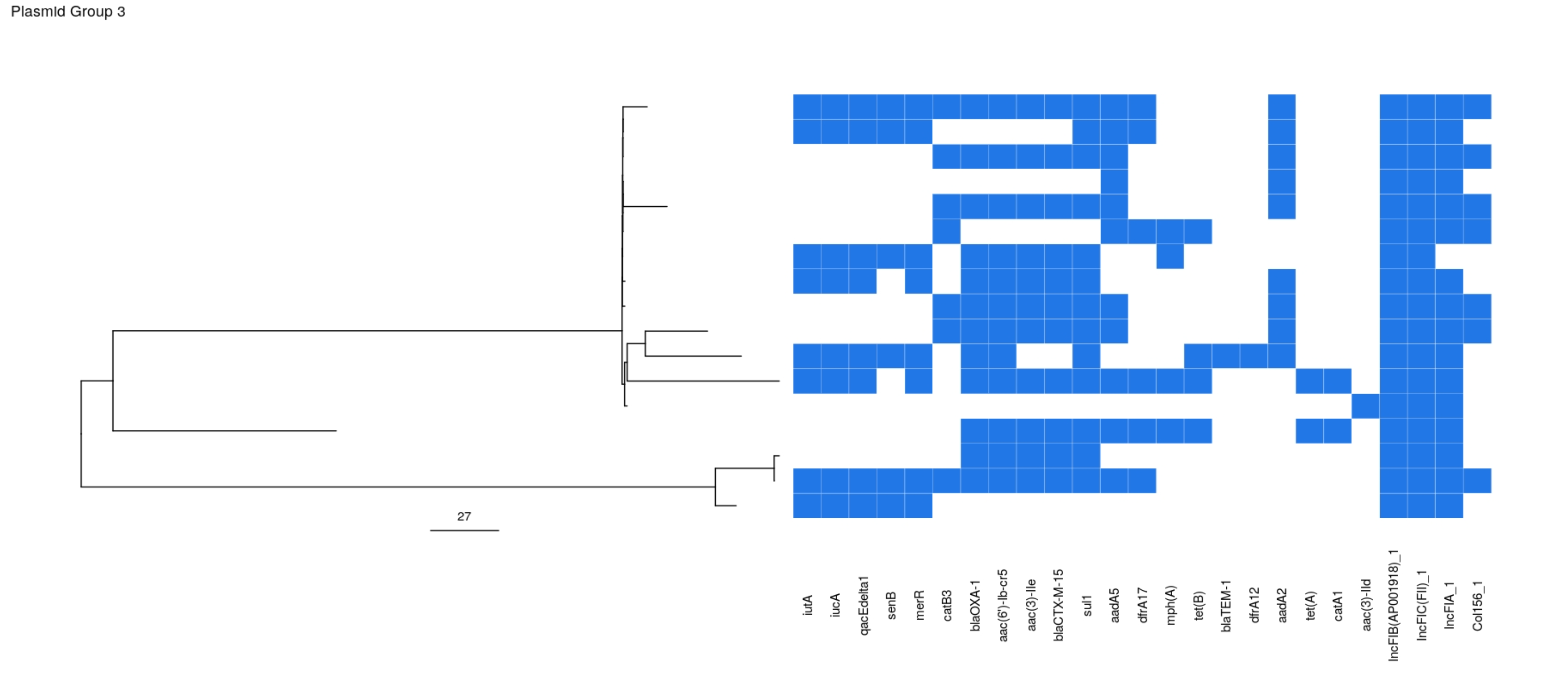

### Fig_S7_new.png

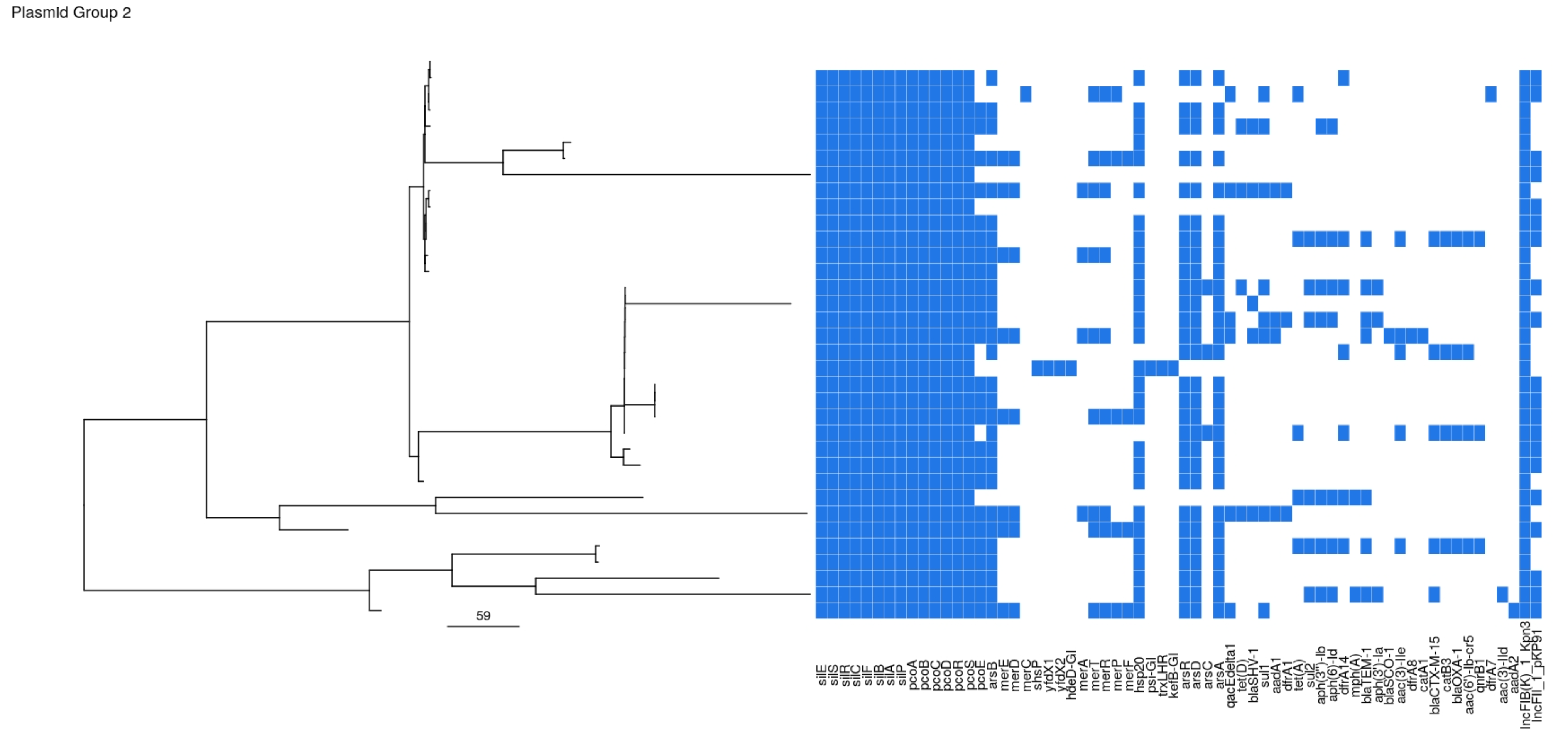

### FigS9_new.png

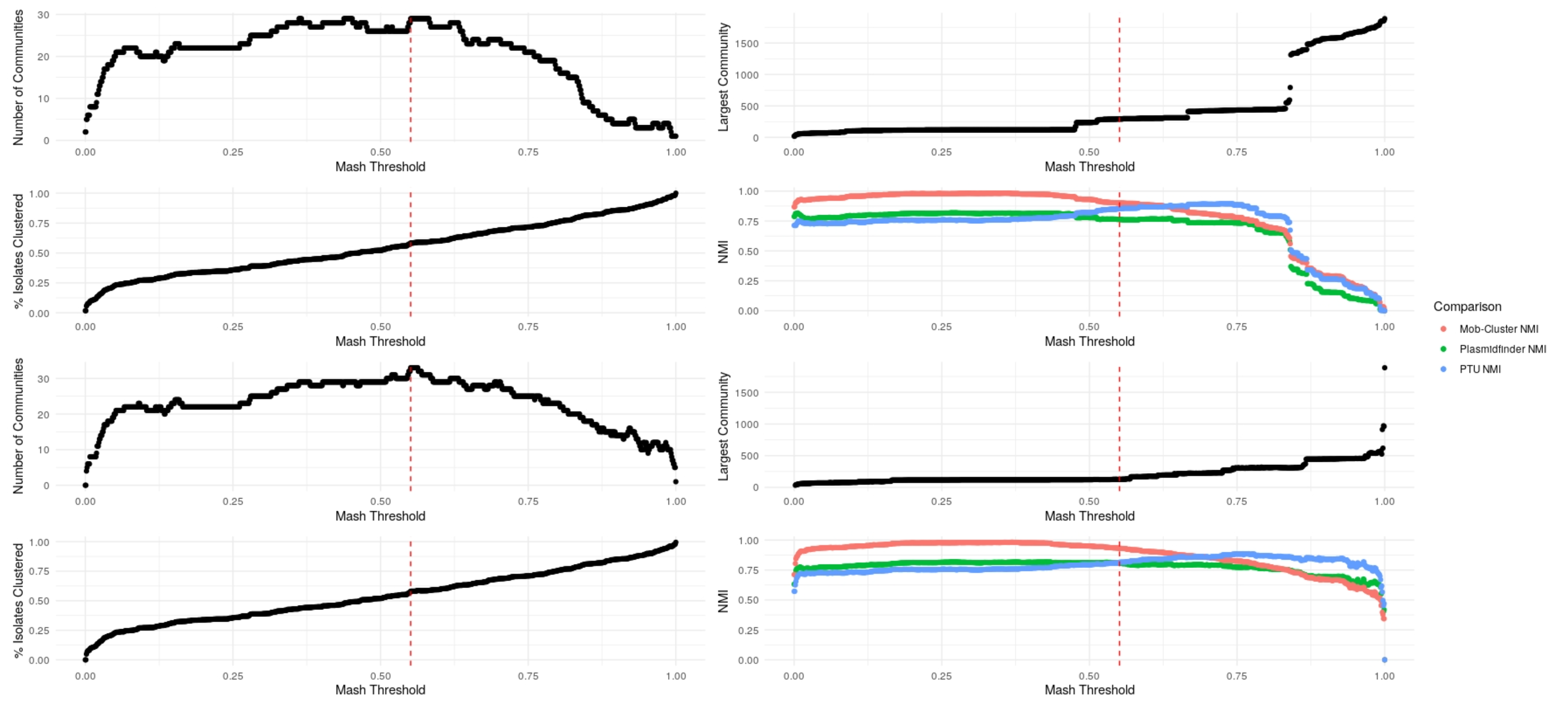

### Figure_S3_new.png

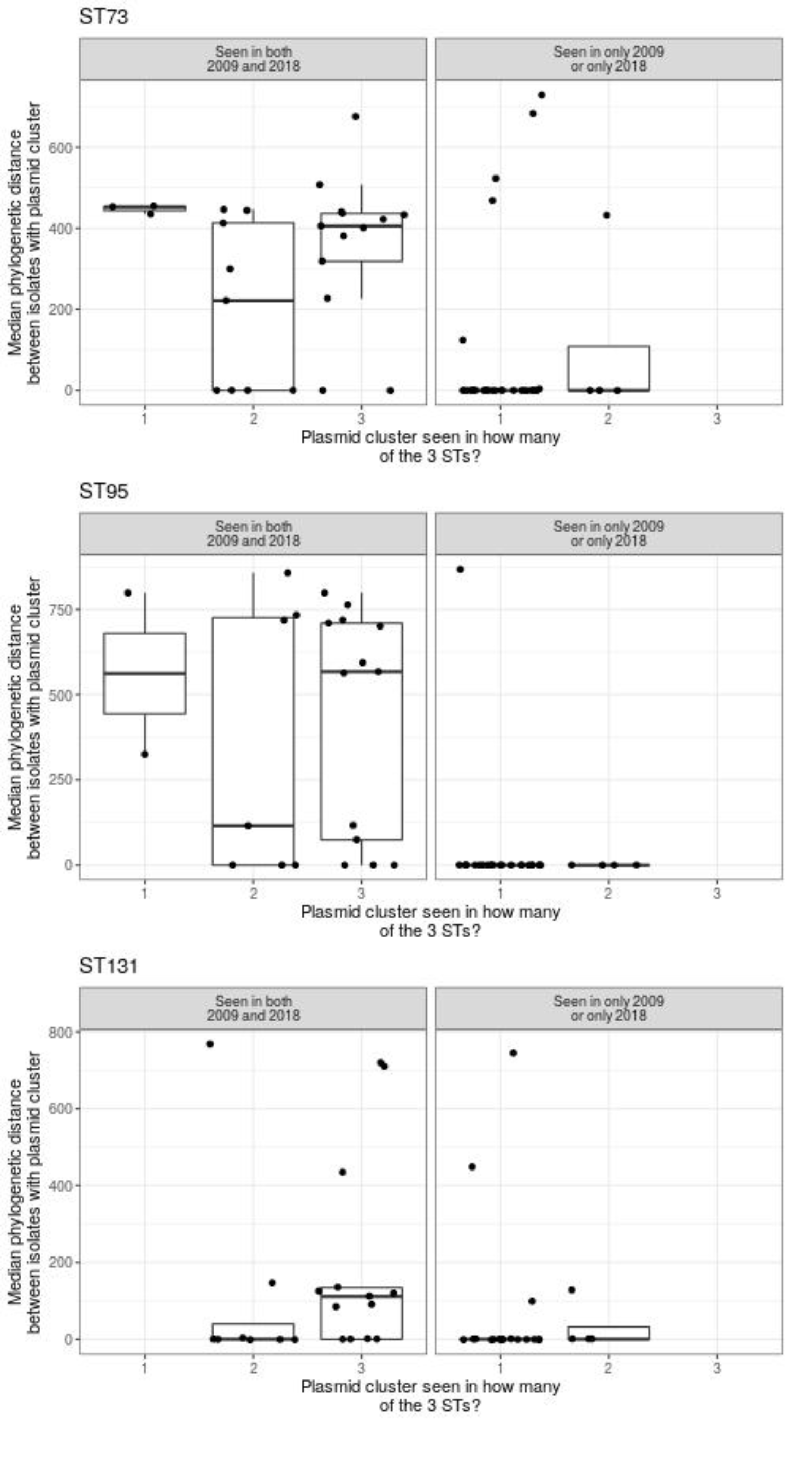

### Figure_S5_new.png

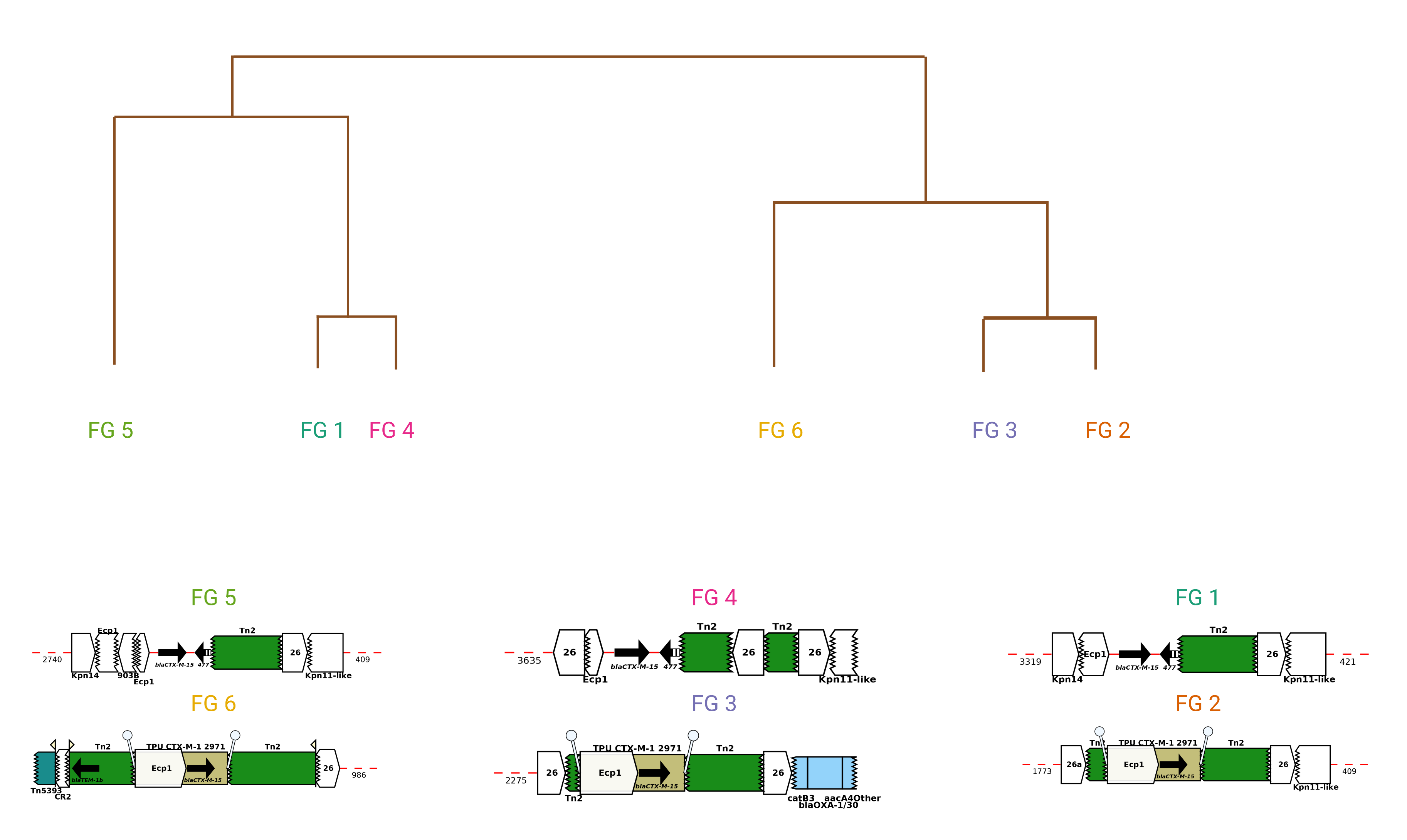
